# Potential health impacts and costs of active case finding guided by *Mycobacterium tuberculosis* immunoreactivity survey results in Blantyre, Malawi

**DOI:** 10.1101/2025.10.01.25337122

**Authors:** Sun Kim, Melike Hazal Can, Hannah M Rickman, Mphatso D Phiri, Marriott Nliwasa, Tisungane E Mwenyenkulu, Kuzani Mbendera, Stéphane Verguet, Marcia C Castro, Elizabeth L Corbett, Peter MacPherson, Ted Cohen, Nicolas A Menzies

## Abstract

**Background:** Active case finding (ACF) for tuberculosis (TB) can reduce transmission, yet efficient targeting requires high-quality surveillance data. We investigated the potential costs and impact of targeted ACF, guided by local estimates of the annual risk of TB infection (ARTI) derived from *Mycobacterium tuberculosis* (*Mtb*) immunoreactivity survey data in children aged <5 years.

**Methods:** Using mathematical models parameterized with local data, we compared three case-finding approaches across 33 urban neighbourhoods in Blantyre, Malawi: passive case finding (PCF) only; PCF with untargeted ACF; and PCF with ARTI-guided targeted ACF. Health outcomes (life expectancy, disability-adjusted life years [DALYs]) and costs were estimated using a Markov microsimulation model. Costs were assessed from health system and societal perspectives. Results were calculated for different assumptions about the relationship between ARTI and true TB prevalence.

**Findings:** Compared to PCF-only, untargeted ACF was estimated to improve life expectancy by 3.7 years (95% credible interval [CrI]: 1.9–5.9) for individuals with TB disease, but at high cost. Targeted ACF covering 48% of the study population was estimated to identify 80% of all individuals with TB and achieved a lower cost per DALY averted (US$1,100, 95% CrI: 900– 1,300) than untargeted ACF (US$1,600, 95% CrI: 1,400–2,000). However, these cost-effectiveness ratios exceeded available cost-effectiveness thresholds for Malawi.

**Interpretation:** Targeting ACF using *Mtb* immunoreactivity survey data could substantially improve health impact and cost-effectiveness compared to untargeted approaches. However, low-cost approaches for collecting these data are needed for wider adoption.

***Funding*:** NIH/NIAID, Wellcome Trust, and NIHR Global Health Research Professorship.

## Introduction

Tuberculosis (TB) remains a major global public health threat, especially in regions with a high burden of HIV.^1^ Active case finding (ACF) is recommended by the World Health Organization to identify individuals with untreated TB, and involves offering systematic screening to community members regardless of symptoms. ACF can be effective in reducing the prevalence of undiagnosed infectious TB, and therefore interrupting community transmission.^2^ While it holds promise, untargeted implementation across entire populations may be expensive and inefficient. Targeting ACF to high prevalence areas may increase efficiency; however, data to identify such areas (e.g., subnational TB prevalence surveys) are usually not available due to the cost and large sample sizes required to conduct such surveys.^3^ TB case notification data could be used as a proxy for prevalence to guide ACF targeting, but areas with high notification rates may reflect capacity for prompt care-seeking behaviour and better access to health services, rather than underlying disease burden. New approaches to prioritising communities with a high burden of undiagnosed disease are required, especially in the context of declining TB burdens.^4^

Malawi is a low-income country in southeastern Africa, with TB incidence of 119 per 100,000 and nearly half of all cases occurring among persons with HIV.^1^ In Blantyre, the second most populous city in the country, high TB notification rates are frequently observed in better-resourced neighbourhoods with stronger health infrastructure, potentially obscuring areas with high levels of undiagnosed TB.^4,5^ Like many high TB burden countries, Malawi’s National Tuberculosis and Leprosy Elimination Program (NTLEP) undertakes ACF, including community-based door-to-door outreach and mobile van-based diagnostic units equipped with Xpert and digital chest X-ray.^6^ But determining which communities should be prioritised for ACF is challenging.

Recent findings from a *Mycobacterium tuberculosis* (*Mtb*) immunoreactivity survey using convenience samples of healthy children under five years of age attending primary care in Blantyre offer an alternative surveillance approach for targeting ACF.^7^ The survey measured *Mtb* immune response using interferon-gamma release assays (IGRAs), providing neighbourhood-level estimates of the annual risk of TB infection (ARTI).^7–10^ ARTI estimates in young children may be particularly informative for identifying areas of recent transmission and elevated force of infection,^11–13^ and thus useful for prioritising locations where undiagnosed TB disease is more likely to be found.

In the present study, we used mathematical modelling to assess the potential health and economic impact of ARTI-guided targeted ACF in Blantyre to inform how surveillance innovations might contribute to more efficient and impactful TB prevention efforts.

## Methods

### Study setting and population

Blantyre is Malawi’s second-largest city and the country’s most densely-populated urban centre, with an estimated population of 800,000.^14^ Adult HIV prevalence is estimated to be 14%, among the highest recorded nationally.^15^ In 2019, the prevalence of TB among adults was estimated at 215 per 100,000, based on a prevalence survey conducted as part of a cluster-randomised trial.^16,17^ A large proportion of Blantyre’s population resides in informal settlements on underdeveloped land, with limited access to health care and other municipal services, despite TB and HIV care being free at the point of delivery.^16,18^

The study focused on 33 urban neighbourhoods (total estimated population of 266,710), defined by community health worker catchment areas and selected based on population density and proximity to 3 study clinics.^19^ Recent neighbourhood ARTI estimates were derived from a *Mtb* immunoreactivity survey, with methods described in detail by Rickman et al. (2025).^7^ These ARTI estimates were used to create scenarios for targeted ACF campaigns, as described below.

### Intervention scenarios

In collaboration with representatives from Malawi’s NTLEP, we defined three hypothetical intervention scenarios. The first scenario, (‘*passive case finding [PCF] only*’), served as a counterfactual baseline with no ACF, in which individuals with TB were only diagnosed if they self-presented to a health facility. The second scenario (*‘untargeted ACF’*) involved PCF in combination with untargeted ACF, implemented uniformly across all clusters regardless of underlying epidemiological differences. The third scenario (*‘targeted ACF’*) involved PCF in combination with targeted ACF, where clusters were prioritized sequentially from highest to lowest ARTI, as estimated from *Mtb* immunoreactivity survey data.

In both ACF scenarios, mobile diagnostic units equipped with digital chest X-ray and Xpert MTB/RIF were deployed to communities, accompanied by community health workers. Community members were invited for screening, beginning with a standardized symptom questionnaire. Individuals reporting TB symptoms (any cough), or with any abnormality on chest X-ray, were asked to provide sputum for testing with Xpert MTB-RIF. Individuals with Xpert-confirmed TB were linked to treatment services. For both ACF scenarios, we calculated results for a one-time community-wide ACF intervention based on a previous randomized trial in Blantyre.^19^ **Figure 1** presents a schematic of the diagnostic pathways and outcomes under each scenario.

**Figure 1:**
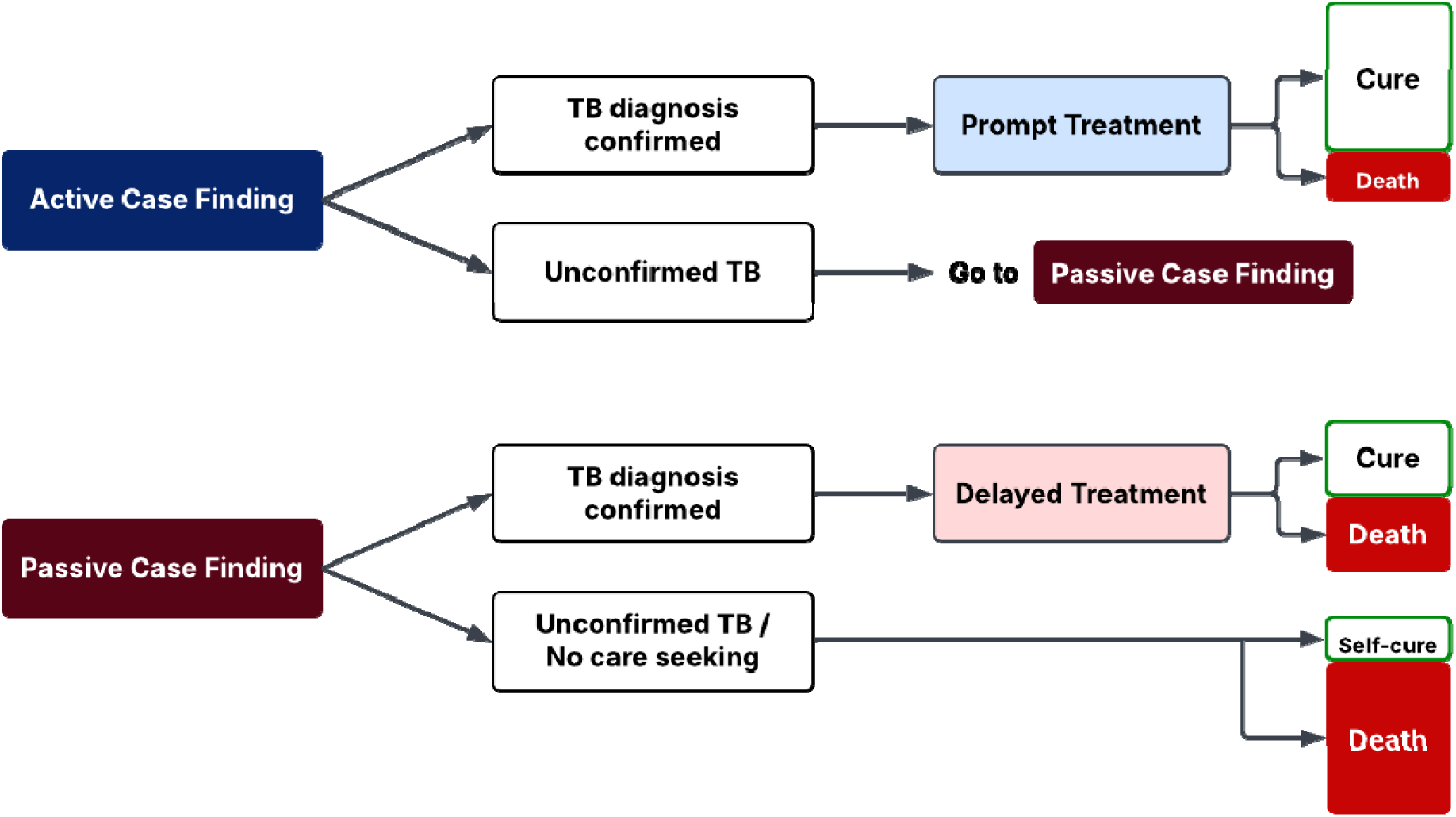
Pathways and outcomes for individuals with TB disease under active and passive case finding.^*^ ^*^The size of each outcome box (cure or death) is illustrative, and reflect relative frequency under each scenario, with larger boxes indicating more common outcomes and smaller boxes indicating less common outcomes.

### Health and cost outcomes

Health outcomes included life expectancy, TB-attributable mortality, and disability-adjusted life years (DALYs). DALYs were calculated as the sum of years of life lost (YLLs) and years lived with disability (YLDs) attributable to TB or HIV. Cost outcomes included both health system expenditures as well as patient-incurred costs, and were expressed in 2022 US dollars (USD). Since TB services are provided free of charge in Malawi, patients were assumed to face no direct medical costs. Direct non-medical costs included transport, food, and accommodation during healthcare visits. Indirect costs reflected income loss or the value of time lost by patients and their households during the TB episode.^20^ For the calculation of incremental cost-effectiveness ratios (ICERs), costs and health benefits (DALYs averted) were both discounted at a rate of 3% per year.

### Modelling approach

We used a Markov simulation model developed by Can et al. (2025)^21^ to estimate lifetime health outcomes and TB care costs for individuals with or without TB and/or HIV under each intervention scenario. The model incorporated transitions between health states, including undiagnosed TB, diagnosis and treatment, post-TB sequelae, and death. Modelled individuals were stratified by TB and HIV status and were simulated under the diagnostic pathways defined in each scenario. **Appendix Table 1** provides the cohort profile used in the model.

Model inputs were drawn from multiple sources, including cluster-level ARTI estimates,^7^ a TB case notification dataset (2022) with residential geolocation collected via tablet-based electronic entry,^4,22,23^ Blantyre TB prevalence survey data (2019),^24^ and population estimates from World Pop (2020).^14^ Cost inputs were based on published literature and NTLEP reports, including unit costs for TB diagnosis, treatment, and HIV-related care, as well as patient-incurred costs (**Appendix Table 2**).

### Sensitivity and specificity of the ACF algorithm

Based on input from the Malawi NTLEP and implementing partners, we assumed that ACF would be undertaken with an algorithm in which individuals with any reported cough would be tested with Xpert. Individuals not reporting a cough would receive chest X-ray, and those with any abnormality on this X-ray would also receive Xpert. Individuals testing Xpert positive would be diagnosed with TB. A 5% pre-treatment loss to follow-up was assumed for individuals diagnosed with TB, based on NTLEP input.

Algorithm sensitivity was calculated as (1-(1-Sens_symp_)*(1-Sens_cxr_))*Sens_xpert_, where Sens_symp_, Sens_cxr_, and Sens_xpert_ represent the sensitivity of the symptom screen, chest X-ray, and Xpert respectively. Algorithm specificity was calculated as 1-(1-Spec_symp_*Spec_cxr_)*(1-Spec_xpert_), where Spec_symp_, Spec_cxr_, and Spec_xpert_ represent the specificity of the symptom screen, chest X-ray, and Xpert respectively. Values and sources for these calculations are shown in **Appendix Table 3**.

### Statistical analysis

Simulation outputs were aggregated at the cluster level to estimate TB disease detection, life expectancy, DALYs, and costs under each intervention scenario. These results were used to calculate incremental differences in health outcomes and costs compared to the *PCF-only* scenario. Incremental cost-effectiveness ratios (ICERs) comparing ACF scenarios to *PCF-only* were computed at both the cluster level and across all clusters combined. The analysis was conducted over a lifetime horizon.

To evaluate the efficiency of ARTI-guided targeted ACF, we assumed that cluster TB prevalence was positively associated with ARTI. We operationalized this by assuming that the TB prevalence in cluster *i* (π_*i*_), is equal to *k* · *A_i_^P^*, where *A_i_* is ARTI in cluster *i*, *P* parameterizes the strength of relationship between ARTI and TB prevalence, and k is a normalizing constant calculated to ensure average prevalence across clusters equals 215 per 100,000: π_*i*_ = *k_i_* · *A_i_^P^*.

We varied the value of *P* from 0.0 (no relationship between TB prevalence and ARTI) to 1.0 (TB prevalence linearly proportional to ARTI), in increments of 0.2. The number of individuals with prevalent TB in each cluster was calculated as π_*i*_ · *N_i_*, where *N_i_* is cluster population.

Using these cluster-specific TB prevalence estimates, we then computed the cumulative number of TB cases identified, DALYs averted, and incremental costs as clusters were sequentially added to the ARTI-guided targeted ACF intervention in order of descending ARTI. To further assess the implications of geographic targeting, we modelled a scenario in which ACF was implemented in clusters comprising approximately half (48%) of the total population (n=128,055), starting with clusters with the highest ARTI. We compared this targeted approach to an untargeted strategy in which the same population coverage was achieved through random selection of clusters.

We computed 95% credible intervals as the 2.5th and 97.5th percentiles of the sample of estimates for each outcome, as generated from 1,000 randomly-drawn parameter sets, reflecting uncertainty in input parameters. All analyses were performed in R software (v4.4.3), using the Rcpp package.^25^

## Results

### Baseline characteristics of clusters

The 33 urban clusters included in the analysis demonstrated substantial variation in population size, TB burden, and HIV prevalence (**Table 1**). Estimates of cluster-specific ARTI ranged from 0 to 9%. **Figure 2** presents the geographic distribution of clusters included in the ARTI survey.

**Table 1:** Baseline characteristics in each cluster.

| Cluster No. | Population | HIV prevalence among TB pop. (%) | *Mtb* immune-reactivity prevalence (%) | ARTI* (%) | TB disease prevalence (per 100,000), under different assumptions for the predictive power of ARTI (*P*) |  |  |  |  |  |
| --- | --- | --- | --- | --- | --- | --- | --- | --- | --- | --- |
|  |  |  |  |  | P =1 | P=0.8 | P=0.6 | P=0.4 | P=0.2 | P=0 |
| 1 | 9537 | 44 | 7.7 | 3.1 | 254 | 262 | 265 | 262 | 255 | 215 |
| 2 | 7675 | 73 | 3.9 | 1.5 | 125 | 149 | 173 | 198 | 221 | 215 |
| 3 | 5897 | 68 | 3.6 | 1.7 | 134 | 157 | 180 | 203 | 224 | 215 |
| 4 | 7524 | 48 | 4.1 | 1.8 | 145 | 167 | 189 | 209 | 228 | 215 |
| 5 | 7198 | 50 | 20 | 8.6 | 701 | 588 | 486 | 394 | 312 | 215 |
| 6 | 5643 | 53 | 0 | 0 | 0 | 0 | 0 | 0 | 0 | 215 |
| 7 | 7837 | 54 | 4.4 | 1.5 | 123 | 146 | 171 | 196 | 220 | 215 |
| 8 | 6573 | 66 | 12 | 5.9 | 480 | 435 | 387 | 338 | 290 | 215 |
| 9 | 8946 | 50 | 3.5 | 1.5 | 118 | 141 | 167 | 193 | 219 | 215 |
| 10 | 11433 | 69 | 3 | 1.2 | 98 | 122 | 149 | 179 | 210 | 215 |
| 11 | 12070 | 65 | 4.7 | 1.8 | 145 | 167 | 189 | 209 | 228 | 215 |
| 12 | 6417 | 45 | 12.1 | 4.6 | 374 | 356 | 333 | 306 | 275 | 215 |
| 13 | 7982 | 65 | 4.7 | 1.9 | 150 | 172 | 193 | 213 | 230 | 215 |
| 14 | 15231 | 48 | 1 | 0.4 | 31 | 48 | 75 | 113 | 167 | 215 |
| 15 | 8054 | 47 | 1.5 | 0.6 | 49 | 70 | 98 | 136 | 183 | 215 |
| 16 | 6476 | 36 | 6.2 | 2.6 | 211 | 225 | 236 | 243 | 245 | 215 |
| 17 | 7904 | 59 | 8.5 | 3.8 | 305 | 302 | 295 | 282 | 264 | 215 |
| 18 | 11002 | 40 | 7 | 3.4 | 275 | 278 | 277 | 271 | 259 | 215 |
| 19 | 7027 | 67 | 15.2 | 6.2 | 503 | 452 | 398 | 345 | 292 | 215 |
| 20 | 3606 | 50 | 14.3 | 5 | 405 | 379 | 350 | 316 | 280 | 215 |
| 21 | 6390 | 61 | 12.8 | 5 | 404 | 379 | 349 | 316 | 280 | 215 |
| 22 | 9435 | 56 | 13.4 | 5.1 | 411 | 384 | 353 | 318 | 281 | 215 |
| 23 | 7233 | 50 | 4.3 | 1.6 | 129 | 152 | 176 | 200 | 223 | 215 |
| 24 | 11967 | 51 | 12.5 | 5.4 | 440 | 405 | 367 | 327 | 284 | 215 |
| 25 | 5369 | 48 | 17.1 | 7.2 | 586 | 510 | 436 | 366 | 301 | 215 |
| 26 | 12976 | 57 | 3.7 | 1.5 | 124 | 148 | 172 | 197 | 221 | 215 |
| 27 | 9715 | 55 | 10.8 | 4.3 | 345 | 334 | 318 | 297 | 271 | 215 |
| 28 | 7038 | 67 | 3.6 | 1.5 | 120 | 144 | 169 | 195 | 219 | 215 |
| 29 | 4463 | 41 | 3.5 | 1.3 | 108 | 132 | 158 | 186 | 215 | 215 |
| 30 | 3934 | 60 | 5.3 | 2 | 164 | 184 | 203 | 220 | 234 | 215 |
| 31 | 6323 | 73 | 0 | 0 | 0 | 0 | 0 | 0 | 0 | 215 |
| 32 | 8574 | 62 | 0 | 0 | 0 | 0 | 0 | 0 | 0 | 215 |
| 33 | 9263 | 69 | 0 | 0 | 0 | 0 | 0 | 0 | 0 | 215 |

\*ARTI refers to the annual risk of tuberculosis infection.

**Figure 2:**
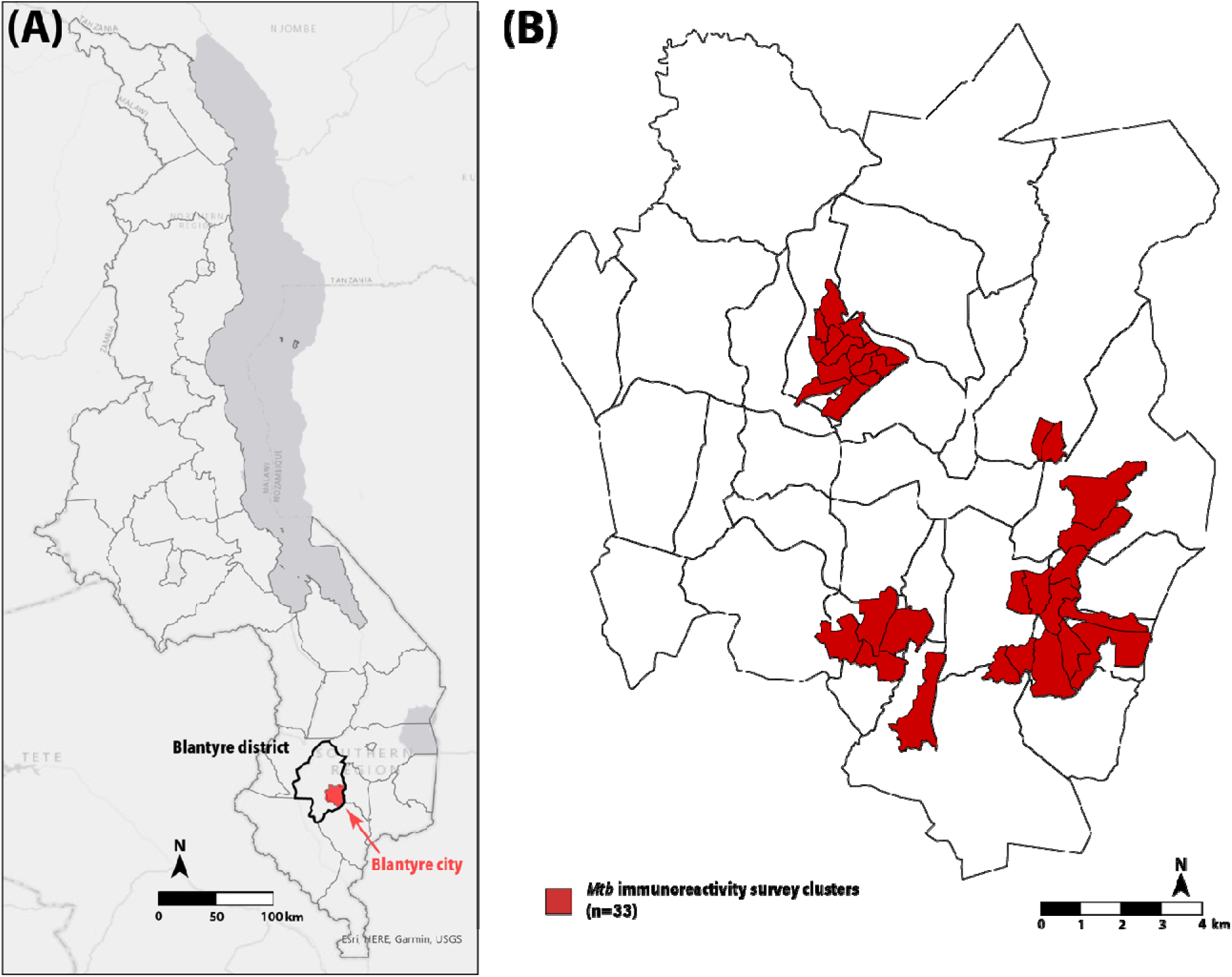
Map of Malawi (Panel A) and Annual Risk of TB Infection (ARTI) by cluster in Blantyre city (Panel B).^*^ ^*^Base map sources: ESRI and https://github.com/petermacp/mlwdata.

### Case detection under varying ARTI predictive power

The number of TB cases identified through ACF varied depending on the assumed predictive power of ARTI. **Figure 3** shows the cumulative number of individuals with TB detected as clusters were sequentially added to the targeted ACF intervention, under predictive power assumptions ranging from *P* = 0 to *P* = 1. When ARTI was assumed to be highly predictive of TB prevalence, a greater proportion of cases was identified in the earliest-added clusters, resulting in a sharply increasing cumulative curve that reflects concentrated yield in a small number of high-ARTI clusters. As predictive power decreased, the distribution of cases became more uniform across clusters, though still yielding greater case detection compared to untargeted ACF.

**Figure 3:**
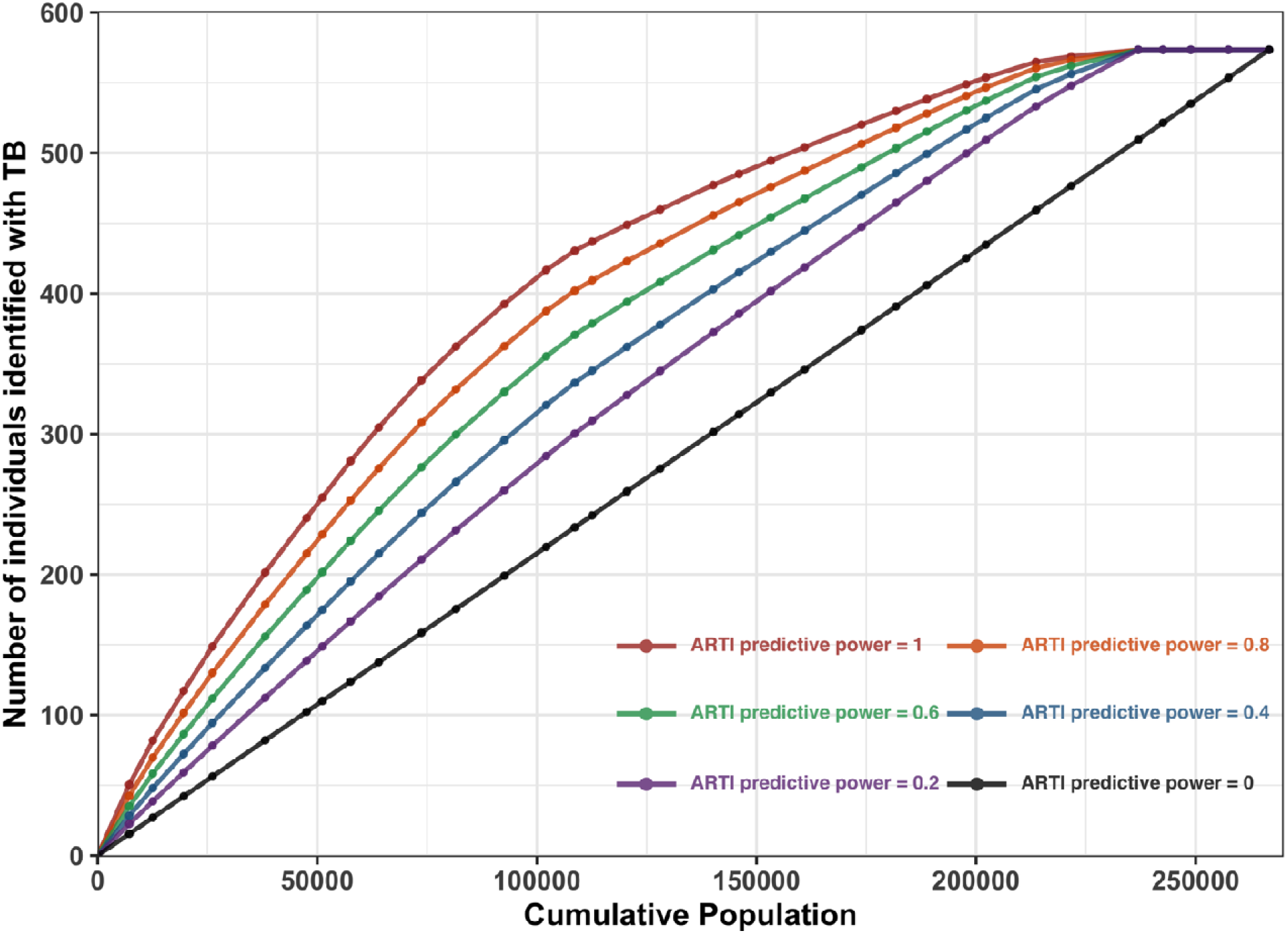
Number of individuals with TB disease identified through active case finding (ACF) based on annual risk of TB infection (ARTI) predictive power.*\** * Excluding participants with false positive results

### Health impact of ACF

ACF implementation resulted in improved health outcomes for individuals with TB. For HIV-negative individuals with TB, ACF increased life expectancy by an average of 3.7 years (95% credible interval [CrI]: 1.9–5.9) compared to PCF alone, while HIV-positive individuals gained an average of 2.3 years (95% CrI: 0.9–4.1) with ACF (**Table 2**). False-positive results for individuals without TB—leading to unnecessary TB treatment—had a limited but non-negligible impact on individuals without TB.

**Table 2:** Estimated life expectancies (LE) under ACF and PCF scenarios.

| Category |  | LE under ACF scenario | LE under PCF scenario | Incremental Difference |
| --- | --- | --- | --- | --- |
| Individuals with TB | HIV-negative | 31.8 (29.8, 33.5) | 28.1 (24.2, 31.5) | 3.7 (1.9, 5.9) |
| Individuals with TB | HIV-positive | 12.8 (6.6, 16.3) | 10.4 (5.3, 14.0) | 2.3 (0.9, 4.1) |
| Individuals without TB | HIV-negative | 37.0 (36.7, 37.3) | 37.0 (36.8, 37.3) | -0.00005<br>(-0.00009, -0.00001) |
| Individuals without TB | HIV-positive | 15.7 (8.0, 20.2) | 15.7 (8.0, 20.2) | -0.00002<br>(-0.00006, 0.00000) |

ACF also led to substantial reductions in DALYs among individuals with TB. Among those who were HIV-negative, the average DALYs averted per person was 6.5 (95% CrI: 3.4–10.2), and among HIV-positive individuals, 3.1 DALYs were averted (95% CrI: 1.2–5.5) (**Table 3**). The majority of these gains were driven by reductions in years of life lost (YLLs) (**Appendix Tables 4** and **5**). Reductions in the share of deaths attributable to TB were also notable: under ACF, the proportion of TB-attributable deaths was 8.1 percentage points lower (95% CrI: 3.7–14.9) among HIV-negative individuals and 15.9 percentage points lower (95% CrI: 8.3–26.4) among HIV-positive individuals, compared with PCF (**Appendix Table 6**).

**Table 3:**
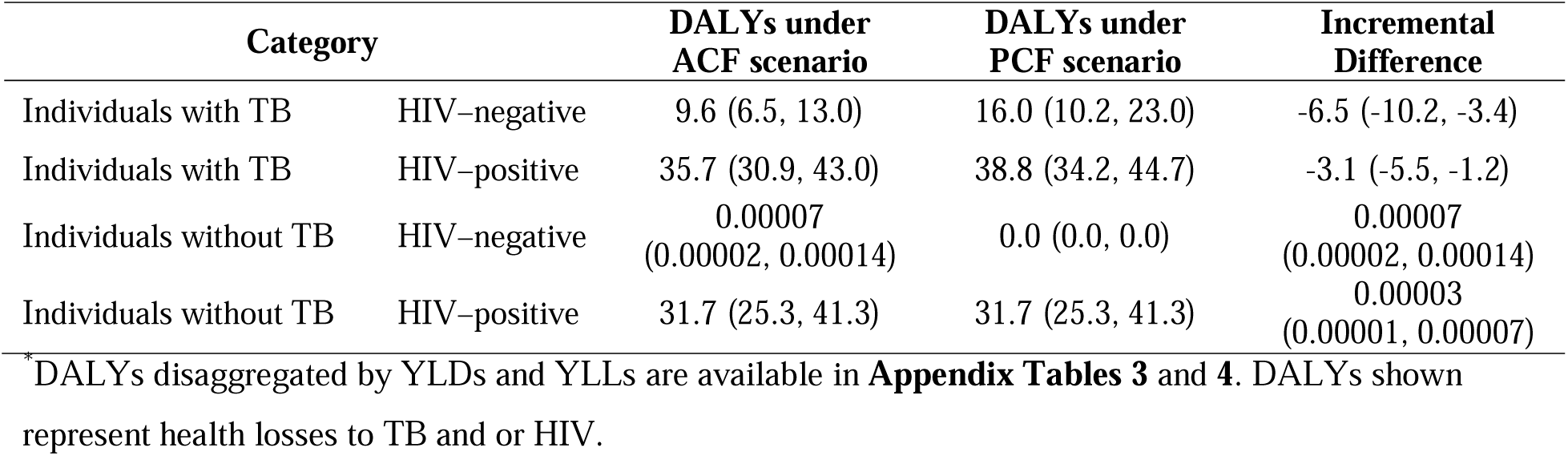
Estimated Disability Adjusted Life Years (DALYs) under ACF and PCF scenarios.

### Cost-effectiveness of ACF by cluster

Cost-effectiveness ratios (comparing ACF to passive detection) varied substantially across clusters, driven by differences in HIV prevalence and estimated TB burden. Under the assumption of perfect predictive power (*P* = 1), the majority of clusters had ICERs below $3,000 per DALY averted, with a median of $2,464 (**Table 4**). As anticipated, the ACF intervention was dominated for clusters with zero ARTI, meaning that for these clusters (with assumed zero TB prevalence) ACF was estimated to incur higher costs and harm health outcomes.

**Table 4:**
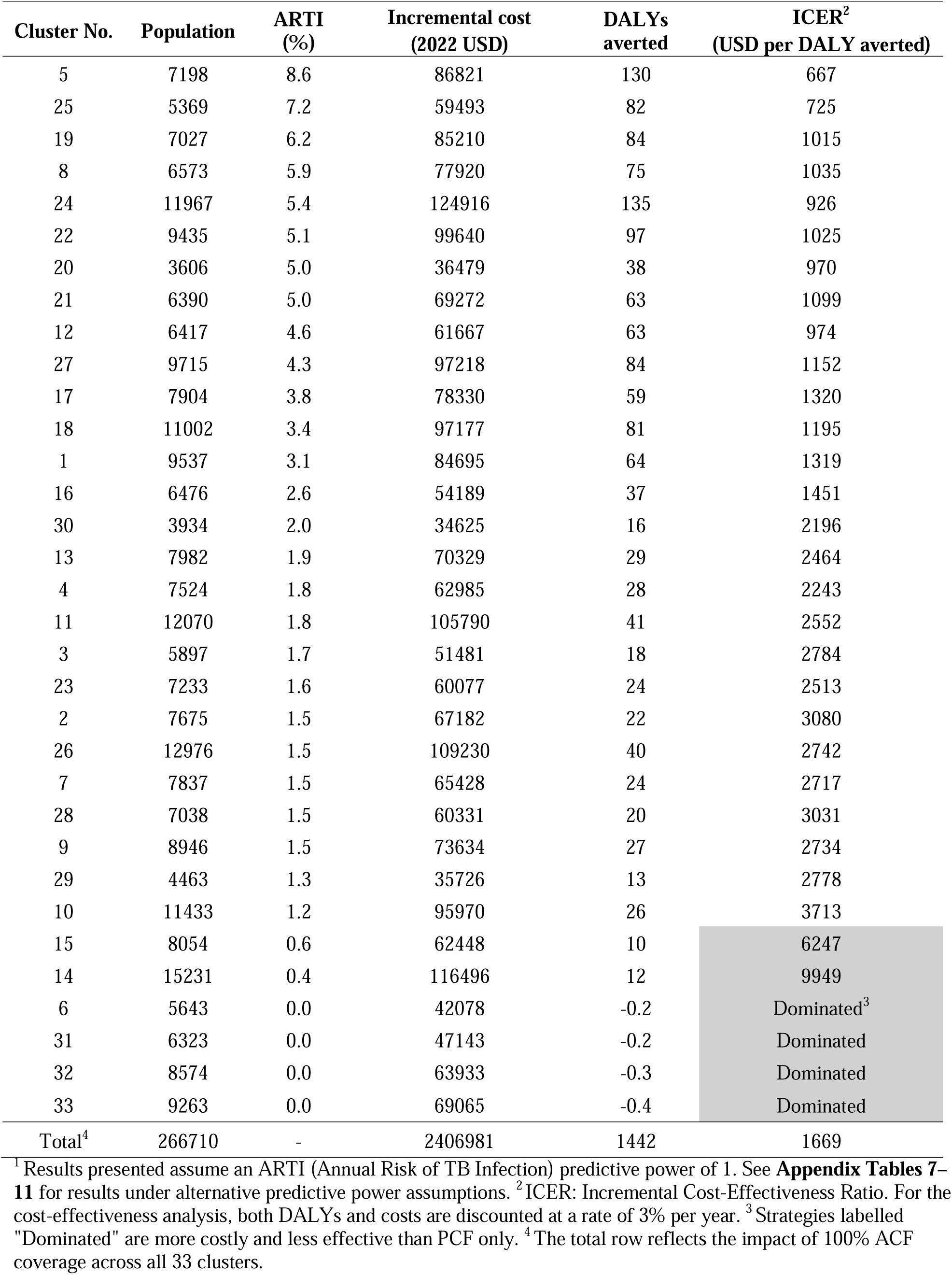
Cost-effectiveness of targeted ACF vs. no ACF by cluster, ordered by ARTI.^1^

| Cluster No. | Population | ARTI (%) | Incremental cost (2022 USD) | DALYs averted | ICER<sup>2</sup> (USD per DALY averted) |
| --- | --- | --- | --- | --- | --- |
| 5 | 7198 | 8.6 | 86821 | 130 | 667 |
| 25 | 5369 | 7.2 | 59493 | 82 | 725 |
| 19 | 7027 | 6.2 | 85210 | 84 | 1015 |
| 8 | 6573 | 5.9 | 77920 | 75 | 1035 |
| 24 | 11967 | 5.4 | 124916 | 135 | 926 |
| 22 | 9435 | 5.1 | 99640 | 97 | 1025 |
| 20 | 3606 | 5.0 | 36479 | 38 | 970 |
| 21 | 6390 | 5.0 | 69272 | 63 | 1099 |
| 12 | 6417 | 4.6 | 61667 | 63 | 974 |
| 27 | 9715 | 4.3 | 97218 | 84 | 1152 |
| 17 | 7904 | 3.8 | 78330 | 59 | 1320 |
| 18 | 11002 | 3.4 | 97177 | 81 | 1195 |
| 1 | 9537 | 3.1 | 84695 | 64 | 1319 |
| 16 | 6476 | 2.6 | 54189 | 37 | 1451 |
| 30 | 3934 | 2.0 | 34625 | 16 | 2196 |
| 13 | 7982 | 1.9 | 70329 | 29 | 2464 |
| 4 | 7524 | 1.8 | 62985 | 28 | 2243 |
| 11 | 12070 | 1.8 | 105790 | 41 | 2552 |
| 3 | 5897 | 1.7 | 51481 | 18 | 2784 |
| 23 | 7233 | 1.6 | 60077 | 24 | 2513 |
| 2 | 7675 | 1.5 | 67182 | 22 | 3080 |
| 26 | 12976 | 1.5 | 109230 | 40 | 2742 |
| 7 | 7837 | 1.5 | 65428 | 24 | 2717 |
| 28 | 7038 | 1.5 | 60331 | 20 | 3031 |
| 9 | 8946 | 1.5 | 73634 | 27 | 2734 |
| 29 | 4463 | 1.3 | 35726 | 13 | 2778 |
| 10 | 11433 | 1.2 | 95970 | 26 | 3713 |
| 15 | 8054 | 0.6 | 62448 | 10 | 6247 |
| 14 | 15231 | 0.4 | 116496 | 12 | 9949 |
| 6 | 5643 | 0.0 | 42078 | -0.2 | Dominated<sup>3</sup> |
| 31 | 6323 | 0.0 | 47143 | -0.2 | Dominated |
| 32 | 8574 | 0.0 | 63933 | -0.3 | Dominated |
| 33 | 9263 | 0.0 | 69065 | -0.4 | Dominated |
| Total<sup>4</sup> | 266710 | - | 2406981 | 1442 | 1669 |

<sup>1</sup> Results presented assume an ARTI (Annual Risk of TB Infection) predictive power of 1. See **Appendix Tables 7–11** for results under alternative predictive power assumptions. <sup>2</sup> ICER: Incremental Cost-Effectiveness Ratio. For the cost-effectiveness analysis, both DALYs and costs are discounted at a rate of 3% per year. <sup>3</sup> Strategies labelled "Dominated" are more costly and less effective than PCF only. <sup>4</sup> The total row reflects the impact of 100% ACF coverage across all 33 clusters.

Figure 4 shows how the total costs and health outcomes of ARTI-guided targeted ACF increased as more clusters were included in the intervention. Under perfect predictive power (*P* = 1), health benefits increased rapidly with initial investments, while as predictive power decreased, the returns to investment diminished, and more clusters were required to achieve the same level of health benefits.

**Figure 4:**
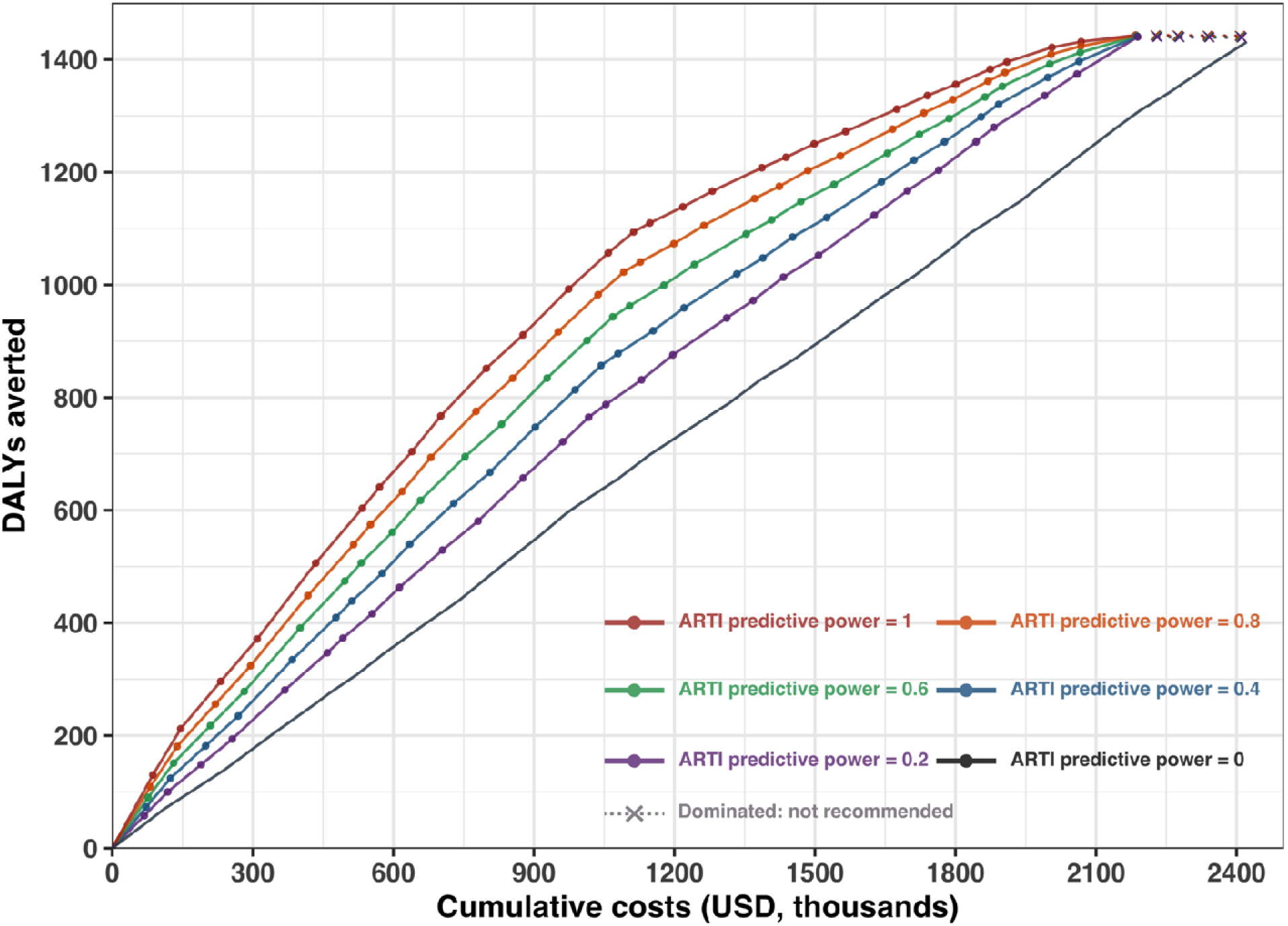
Cumulative costs and health benefits of expanding ACF under varying ARTI predictive power assumptions. *Dominated points (marked ×) represent clusters where ACF was more costly and less effective than no ACF.

### Comparison of targeted vs. untargeted ACF

As shown in **Table 5**, a targeted ACF intervention including 48% of the total population consistently identified clusters with higher average TB prevalence, ranging from 362 to 269 per 100,000 as predictive power decreased from *P* = 1 to *P* = 0.2. This targeting approach captured a larger share of the total TB burden than untargeted ACF. For example, when ARTI was assumed to be perfectly predictive (*P* = 1), targeting 48% of the population using prioritized clusters identified 80% of all TB cases potentially identifiable through ACF, compared to only 48% in the untargeted scenario.

**Table 5:** Cost-effectiveness of targeted ACF vs. untargeted ACF at 48% cluster coverage.^1^

|  | Average TB<br>prevalence among<br>selected clusters<br>(per 100k) | Fraction of<br>total cases<br>identified (%) | Incremental cost<sup>2</sup><br>in thousand | DALYs averted | ICER<br>compared to<br>no ACF |
| --- | --- | --- | --- | --- | --- |
| **Targeted**<br>**(*P*=1)** | 362 | 80.2 | 1281<br>(761, 1848) | 1166<br>(812, 1425) | 1098<br>(937, 1296) |
| **Targeted**<br>**(*P*=0.8)** | 342 | 76.0 | 1263<br>(750, 1824) | 1105<br>(769, 1352) | 1143<br>(976, 1349) |
| **Targeted**<br>**(*P*=0.6)** | 320 | 71.2 | 1243<br>(739, 1798) | 1036<br>(720, 1268) | 1199<br>(1025, 1418) |
| **Targeted**<br>**(*P*=0.4)** | 295 | 65.9 | 1221<br>(726, 1769) | 959<br>(667, 1175) | 1272<br>(1089, 1505) |
| **Targeted**<br>**(*P*=0.2)** | 269 | 60.1 | 1197<br>(712, 1738) | 876<br>(608, 1073) | 1367<br>(1170, 1619) |
| **Untargeted**<br>**(*P*=0)** | 215 | 48.0 | 1148<br>(683, 1673) | 699<br>(485, 857) | 1643<br>(1408, 1953) |

<sup>1</sup> For the cost-effectiveness analysis, both DALYs and costs (in 2022 USD) are discounted at a rate of 3% per year.<sup>2</sup> Incremental cost is assessed relative to the no-ACF scenario.

As predictive power decreased from 1 to 0.2, ICERs increased from $1,100 (95% CrI: 900–1,300) to $1,400 (95% CrI: 1,200–1,600) per DALY averted. In contrast, untargeted ACF (*P* = 0) produced an ICER of $1,600 (95% CrI: 1,400–2,000) per DALY averted.

## Discussion

This study shows that geographically-targeted ACF using ARTI estimates derived from a novel convenience sampling approach for *Mtb* immunoreactivity in young children can result in greater health gains and efficiency compared to untargeted ACF. In Blantyre, Malawi—a setting with a high burden of TB and HIV—we found that differences in community-level ARTI, as measured through *Mtb* immunoreactivity surveys, could help prioritize areas for diagnostic interventions more effectively than untargeted approaches. Combining novel approaches to TB surveillance (convenience *Mtb* immunoreactivity sampling among under 5-year-old children) with targeted ACF therefore offers a new approach to increase the effectiveness and efficiency of TB screening in high burden settings.

Prioritising where to implement ACF has been a long-standing challenge for TB programmes.^3^ Although modelling analyses have suggested that targeted ACF could offer substantial efficiency gains in reducing transmission,^26^ systematic reviews of implementation studies have identified only a small number of studies that evaluated the effectiveness of targeted TB interventions,^27,28^ likely because of the difficulty in identifying a suitable epidemiological marker to guide targeting. Our findings suggest that ARTI data could fill this gap, offering a feasible and practical epidemiological marker for directing ACF implementation.

For programs planning to implement ACF, how to best deploy ACF services is a central question. In our analysis, compared to PCF alone, ACF improved life expectancy, reduced TB-attributable mortality, and averted approximately 553 DALYs per 100,000 individuals screened, on average across scenarios. Efficiency gains were observed when comparing targeted to untargeted ACF.

For example, providing ACF to approximately half of the population through targeted ACF could find 80% of all TB cases potentially identifiable through ACF, with more favourable ICERs compared to untargeted ACF. These findings highlight the potential value of geographic targeting in enhancing the efficiency of ACF implementation under real-world constraints on population coverage.

While targeted ACF improved efficiency relative to untargeted ACF, the absolute cost-effectiveness of the intervention remains a critical consideration.^29,30^ For Malawi, published estimates suggest that cost-effectiveness threshold may lie between 33% and 44% of per-capita GDP (equivalent to $199 to $265 in 2022 USD).^31^ A recent government report proposed a more stringent threshold of $66 per DALY averted.^32^ By these benchmarks, the ICERs estimated for both targeted and untargeted ACF—ranging from $1,100 to $1,600 per DALY averted—would not meet conventional definitions of cost-effectiveness, and represent a substantial cost relative to available healthcare funding Malawi, where per capita health expenditure was approximately US$40 in 2022. However, for infectious diseases like TB such thresholds may underestimate longer-term population benefits, including transmission reduction, mitigation of future risks of drug-resistant disease, and progress toward elimination goals. Moreover, the cost-effectiveness of ACF could improve under settings with higher TB prevalence or with reduced implementation costs.

These findings suggest that although ACF is a high-cost intervention, its efficiency can be substantially improved through geographic prioritization, potentially offering a higher return on investment in settings where ACF is already being implemented—as is the case in Blantyre and across Malawi. Although this study used ARTI estimates to guide targeted ACF, the modelling framework is data-source agnostic and can be parameterized using alternative proxies for TB burden, including case notification rates, genomic risk predictions, or community prevalence estimates. Each of these other data sources representing alternative possible approaches for targeting ACF.

A key contribution of this study lies in demonstrating the potential of using *Mtb* immunoreactivity survey data as a basis for targeting ACF. In many high-burden urban settings, including Blantyre, TB notification data may not accurately reflect underlying transmission risk due to variations in health system access, diagnostic capacity, or health-seeking behavior.^4^ Historically, ARTI data have been widely used to track TB epidemiology and inform public health policy and planning, including prioritization of interventions such as case finding, preventive therapy, and vaccination strategies.^33–37^ While resource requirements exist, immunoreactivity surveys are considerably less intensive than TB prevalence surveys and can be feasibly integrated into existing surveillance systems, particularly within primary care settings.^38^ Accordingly, the cost of survey implementation was excluded from our analysis, as the data were collected through a separate research study that was embedded within primary care visits, and not as part of the intervention being evaluated.

Our model projected that ACF substantially reduced TB-attributable deaths, thereby increasing life expectancy among both people with and without HIV, underscoring its potential value in mitigating excess TB-associated mortality. This contrasts with findings from the TREATS study in Zambia and South Africa, which found no significant reduction in TB prevalence following a community-based intervention combining HIV testing, linkage to care, and TB symptom screening.^39^ However, unlike TREATS, our model did not include dynamic transmission effects, and therefore does not capture potential population-level benefits of ACF in interrupting TB transmission. The observed differences may also reflect variation in screening approaches: TREATS primarily relied on symptom screening, whereas our ACF strategy also used chest X-ray with computer-aided detection for individuals without symptoms. Furthermore, ACF may mitigate long-term morbidity associated with post-TB sequelae, which are increasingly recognized as major contributors to TB disease burden.^21,40^ These sequelae are often underdiagnosed and underreported yet contribute significantly to the burden of DALYs in high TB-burden settings.^21^ By allowing earlier identification and initiation of treatment, ACF may prevent progression to severe TB disease and reduce the likelihood of permanent impairment.

At the cluster level, we observed substantial variation in the economic efficiency of ACF, with high-ARTI clusters generally achieving lower ICERs. These findings support a phased approach to ACF scale-up: initiating ACF in high-ARTI clusters and expanding outward as resources allow. However, interpretation of value for money should consider not only cost per DALY but also health system priorities, budget constraints, and equity considerations, particularly in resource-limited settings.^3,30^

This study has several limitations. First, although our simulations assumed a correlation between ARTI and active TB disease prevalence, the strength of this relationship may vary depending on factors such as age, immune status, test performance, and sampling uncertainty. In addition, ARTI estimates themselves are subject to uncertainty, particularly in clusters with zero positives, which may reflect limited sample size rather than true absence of infection. To explore this, we modelled a range of predictive power scenarios; nonetheless, further empirical validation of the link between ARTI and TB prevalence is needed.^37,41,42^ Second, our analysis focused on direct health outcomes and costs and did not capture potential benefits from reduced secondary transmission. If secondary transmission effects were uniformly distributed across clusters— effectively acting as a multiplier on the health gains associated with identifying each prevalent TB case—the relative advantage of targeting would remain unchanged, but the overall ICERs would improve proportionally. In this case, ACF would appear more cost-effective in absolute terms, though the comparative efficiency of targeted versus untargeted strategies would remain constant. Third, we examined a scenario in which immunoreactivity survey data were already available from an earlier study. In other settings the costs of these surveys would need to be considered, in addition to options for making them more efficient.

Evidence describing even modest differences in transmission can inform more strategic and efficient ACF delivery. Building on data sources like convenience-based immunoreactivity surveys, this approach can be embedded in national surveillance systems and adapted to other high-burden settings. While targeted ACF improves efficiency, it raises equity concerns and cost implications—particularly around prioritization and long-term ART provision following extended survival. These downstream costs, though linked to positive outcomes, should be considered in programmatic planning.

In conclusion, geographically-targeted TB ACF directed by ARTI data from convenience surveys of MTB immunoreactivity offers a data-driven strategy to improve the efficacy and effectiveness of community-based TB screening. Even in settings with relatively uniform burden, immunoreactivity data can support more targeted resource allocation and strengthen the case for scaling up ACF. Embedding targeted approaches within existing systems—while considering equity and long-term costs—may improve the effectiveness and sustainability of TB programs.

**Contributors** (Contributor Role Taxonomy (CRediT) table: https://credit.niso.org) Conceptualization: SK, PM, SV, MCC, TC, NAM; Data curation: SK, MHC, HMR, PM; Formal analysis: SK, MHC; Funding acquisition: ELC, PM, TC, NAM; Investigation: SK, PM, TC, NAM; Methodology: SK, MHC; Project administration: SK; Resources: HMR, MDP, MN, TEM, KM, ELC, PM; Software: SK, MHC; Supervision: PM, SV, MCC, TC, NM; Validation: PM, TC, NAM; Visualization: SK; Writing–original draft: SK; Writing–review & editing: all authors.

## Data Availability

All data produced in the present study are available upon reasonable request to the authors.

## Acknowledgements

PM was funded by Wellcome (304666/Z/23/Z) and an NIHR Global Health Research Professorship (NIHR304311). The views expressed are those of the author(s) and not necessarily those of the NIHR or the Department of Health and Social Care. For the purpose of open access, the author has applied a CC BY public copyright licence to any Author Accepted Manuscript version arising from this submission. MP was funded by the UK Foreign, Commonwealth and Development Office [“Leaving no-one behind: transforming gendered pathways to health for TB”; (2018/S 196-443482]; however, the views expressed do not necessarily reflect the UK government’s official policies.

## Appendices

**Appendix Table 1:** Cohort profile for simulation model.*

|  | Malawi analytic cohort |
| --- | --- |
| Number of individuals | 10,000 |
| Male (%) | 75.0 |
| Age (mean, IQR) | 34.0 (26.0 – 40.0) |
| Living with HIV (%) | 13.6 |
| Receiving ART, among persons with HIV (%) | 70.1 |
| CD4 cell count (cells/μL; mean, IQR) | 274 (68 – 372) |

\*A full list of simulation model parameters and model details is provided in Can et al. (2025)<sup>21</sup>

**Appendix Table 2:**
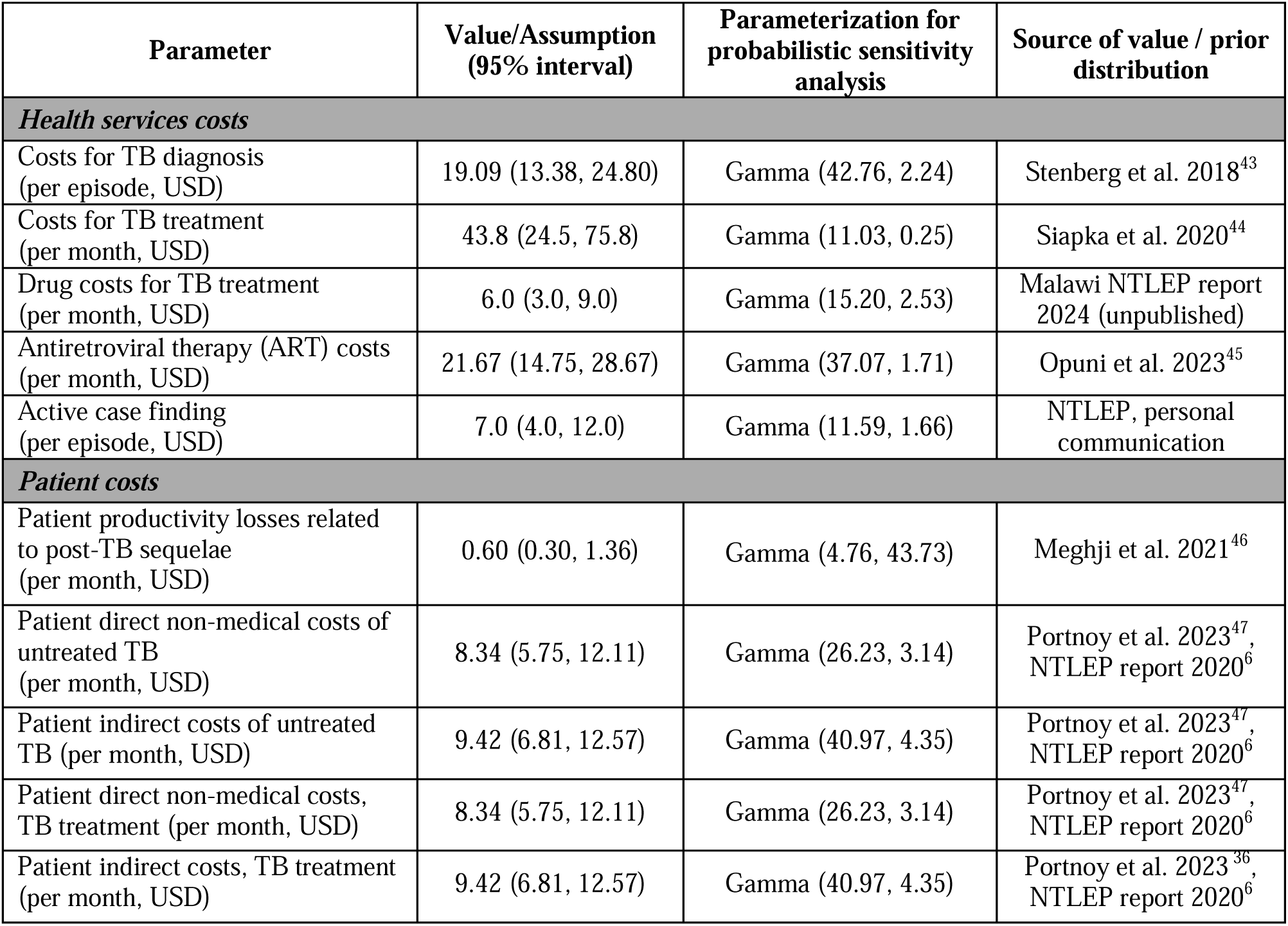
Cost parameter inputs.

| Parameter | Value/Assumption<br>(95% interval) | Parameterization for<br>probabilistic sensitivity<br>analysis | Source of value / prior<br>distribution |
| --- | --- | --- | --- |
| **Health services costs** |  |  |  |
| Costs for TB diagnosis<br>(per episode, USD) | 19.09 (13.38, 24.80) | Gamma (42.76, 2.24) | Stenberg et al. 2018<sup>43</sup> |
| Costs for TB treatment<br>(per month, USD) | 43.8 (24.5, 75.8) | Gamma (11.03, 0.25) | Siapka et al. 2020<sup>44</sup> |
| Drug costs for TB treatment<br>(per month, USD) | 6.0 (3.0, 9.0) | Gamma (15.20, 2.53) | Malawi NTLEP report<br>2024 (unpublished) |
| Antiretroviral therapy (ART) costs<br>(per month, USD) | 21.67 (14.75, 28.67) | Gamma (37.07, 1.71) | Opuni et al. 2023<sup>45</sup> |
| Active case finding<br>(per episode, USD) | 7.0 (4.0, 12.0) | Gamma (11.59, 1.66) | NTLEP, personal<br>communication |
| **Patient costs** |  |  |  |
| Patient productivity losses related<br>to post-TB sequelae<br>(per month, USD) | 0.60 (0.30, 1.36) | Gamma (4.76, 43.73) | Meghji et al. 2021<sup>46</sup> |
| Patient direct non-medical costs of<br>untreated TB<br>(per month, USD) | 8.34 (5.75, 12.11) | Gamma (26.23, 3.14) | Portnoy et al. 2023<sup>47</sup>,<br>NTLEP report 2020<sup>6</sup> |
| Patient indirect costs of untreated<br>TB (per month, USD) | 9.42 (6.81, 12.57) | Gamma (40.97, 4.35) | Portnoy et al. 2023<sup>47</sup>,<br>NTLEP report 2020<sup>6</sup> |
| Patient direct non-medical costs,<br>TB treatment (per month, USD) | 8.34 (5.75, 12.11) | Gamma (26.23, 3.14) | Portnoy et al. 2023<sup>47</sup>,<br>NTLEP report 2020<sup>6</sup> |
| Patient indirect costs, TB treatment<br>(per month, USD) | 9.42 (6.81, 12.57) | Gamma (40.97, 4.35) | Portnoy et al. 2023<sup>36</sup>,<br>NTLEP report 2020<sup>6</sup> |

**Appendix Table 3:** Sensitivity and specificity values for the ACF algorithm

|  | Sensitivity | Specificity | Sources |
| --- | --- | --- | --- |
| Symptom screen (any cough) | 42.0% | 94.0% | World Health Organization, 2021<sup>48</sup> |
| Chest X-ray (any abnormality) | 94.0% | 89.0% | World Health Organization, 2021<sup>48</sup> |
| Xpert | 84.7% | 99.8% | Zifodya et al. 2021<sup>49</sup>, Veeken et al. 2025<sup>50</sup> |
| Algorithm | 82.21% | 99.96% | Calculated |

**Appendix Table 4:**
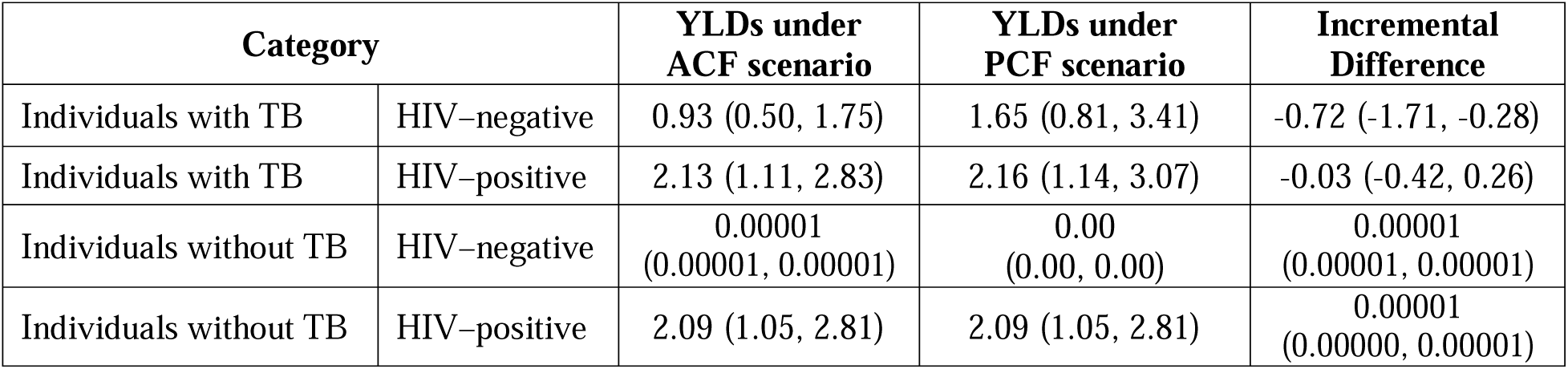
Estimated years of healthy life lost due to disability (YLDs).

| Category |  | YLDs under ACF scenario | YLDs under PCF scenario | Incremental Difference |
| --- | --- | --- | --- | --- |
| Individuals with TB | HIV–negative | 0.93 (0.50, 1.75) | 1.65 (0.81, 3.41) | -0.72 (-1.71, -0.28) |
| Individuals with TB | HIV–positive | 2.13 (1.11, 2.83) | 2.16 (1.14, 3.07) | -0.03 (-0.42, 0.26) |
| Individuals without TB | HIV–negative | 0.00001<br>(0.00001, 0.00001) | 0.00<br>(0.00, 0.00) | 0.00001<br>(0.00001, 0.00001) |
| Individuals without TB | HIV–positive | 2.09 (1.05, 2.81) | 2.09 (1.05, 2.81) | 0.00001<br>(0.00000, 0.00001) |

**Appendix Table 5:** Estimated years of life lost due to premature mortality (YLLs).

| Category |  | YLLs under ACF scenario | YLLs under PCF scenario | Incremental Difference |
| --- | --- | --- | --- | --- |
| Individuals with TB | HIV–negative | 8.64 (5.89, 11.62) | 14.39 (9.33, 20.27) | -5.76 (-9.16, -3.12) |
| Individuals with TB | HIV–positive | 33.6 (28.5, 41.7) | 36.7 (31.8, 43.3) | -3.08 (-5.50, -1.11) |
| Individuals without TB | HIV–negative | 0.00007<br>(0.00002, 0.00014) | 0.00<br>(0.00, 0.00) | 0.00007<br>(0.00002, 0.00014) |
| Individuals without TB | HIV–positive | 29.6 (23.2, 40.1) | 29.6 (23.2, 40.1) | 0.00002<br>(0.00000, 0.00007) |

**Appendix Table 6:** Estimated TB-attributable deaths (as a percentage of total deaths, %).

| Category |  | Deaths under ACF scenario | Deaths under PCF scenario | Incremental Difference |
| --- | --- | --- | --- | --- |
| Individuals with TB | HIV–negative | 8.1 (4.7, 13.2) | 16.2 (8.5, 27.7) | -8.1 (-14.9, -3.7) |
| Individuals with TB | HIV–positive | 20.6 (13.4, 30.1) | 36.5 (22.6, 55.6) | -15.9 (-26.4, -8.3) |

**Appendix Table 7:** Cost-effectiveness of targeted ACF by cluster (*P*\*=0.8).

| Cluster No. | Population | ARTI | Incremental cost (2022 USD) | DALYs averted | ICER (USD per DALY averted) |
| --- | --- | --- | --- | --- | --- |
| 5 | 7198 | 0.0862 | 81514 | 109 | 746 |
| 25 | 5369 | 0.0721 | 56971 | 71 | 797 |
| 19 | 7027 | 0.0619 | 81844 | 75 | 1087 |
| 8 | 6573 | 0.0591 | 75196 | 68 | 1103 |
| 24 | 11967 | 0.0541 | 122130 | 124 | 983 |
| 22 | 9435 | 0.0506 | 97717 | 91 | 1077 |
| 20 | 3606 | 0.0498 | 35878 | 35 | 1018 |
| 21 | 6390 | 0.0497 | 67924 | 59 | 1150 |
| 12 | 6417 | 0.046 | 61008 | 60 | 1012 |
| 27 | 9715 | 0.0425 | 96413 | 82 | 1181 |
| 17 | 7904 | 0.0375 | 78176 | 59 | 1328 |
| 18 | 11002 | 0.0338 | 97371 | 82 | 1182 |
| 1 | 9537 | 0.0313 | 85083 | 66 | 1289 |
| 16 | 6476 | 0.0259 | 54592 | 40 | 1368 |
| 30 | 3934 | 0.0202 | 35274 | 18 | 1991 |
| 13 | 7982 | 0.0185 | 71871 | 33 | 2201 |
| 4 | 7524 | 0.0178 | 64028 | 32 | 1978 |
| 11 | 12070 | 0.0178 | 108182 | 48 | 2263 |
| 3 | 5897 | 0.0165 | 52751 | 22 | 2436 |
| 23 | 7233 | 0.0159 | 61169 | 28 | 2169 |
| 2 | 7675 | 0.0154 | 69027 | 26 | 2664 |
| 26 | 12976 | 0.0153 | 111562 | 47 | 2355 |
| 7 | 7837 | 0.0151 | 66756 | 29 | 2325 |
| 28 | 7038 | 0.0148 | 61862 | 24 | 2596 |
| 9 | 8946 | 0.0145 | 75018 | 32 | 2317 |
| 29 | 4463 | 0.0133 | 36266 | 16 | 2305 |
| 10 | 11433 | 0.012 | 98608 | 32 | 3052 |
| 15 | 8054 | 0.006 | 63481 | 14 | 4397 |
| 14 | 15231 | 0.0038 | 118163 | 19 | 6322 |
| 6 | 5643 | 0 | 42078 | -0.2 | Dominated |
| 31 | 6323 | 0 | 47143 | -0.2 | Dominated |
| 32 | 8574 | 0 | 63933 | -0.3 | Dominated |
| 33 | 9263 | 0 | 69065 | -0.4 | Dominated |
| Total | 266710 | - | 2408054 | 1441 | 1671 |

\* $P$ denotes predictive power

**Appendix Table 8:**
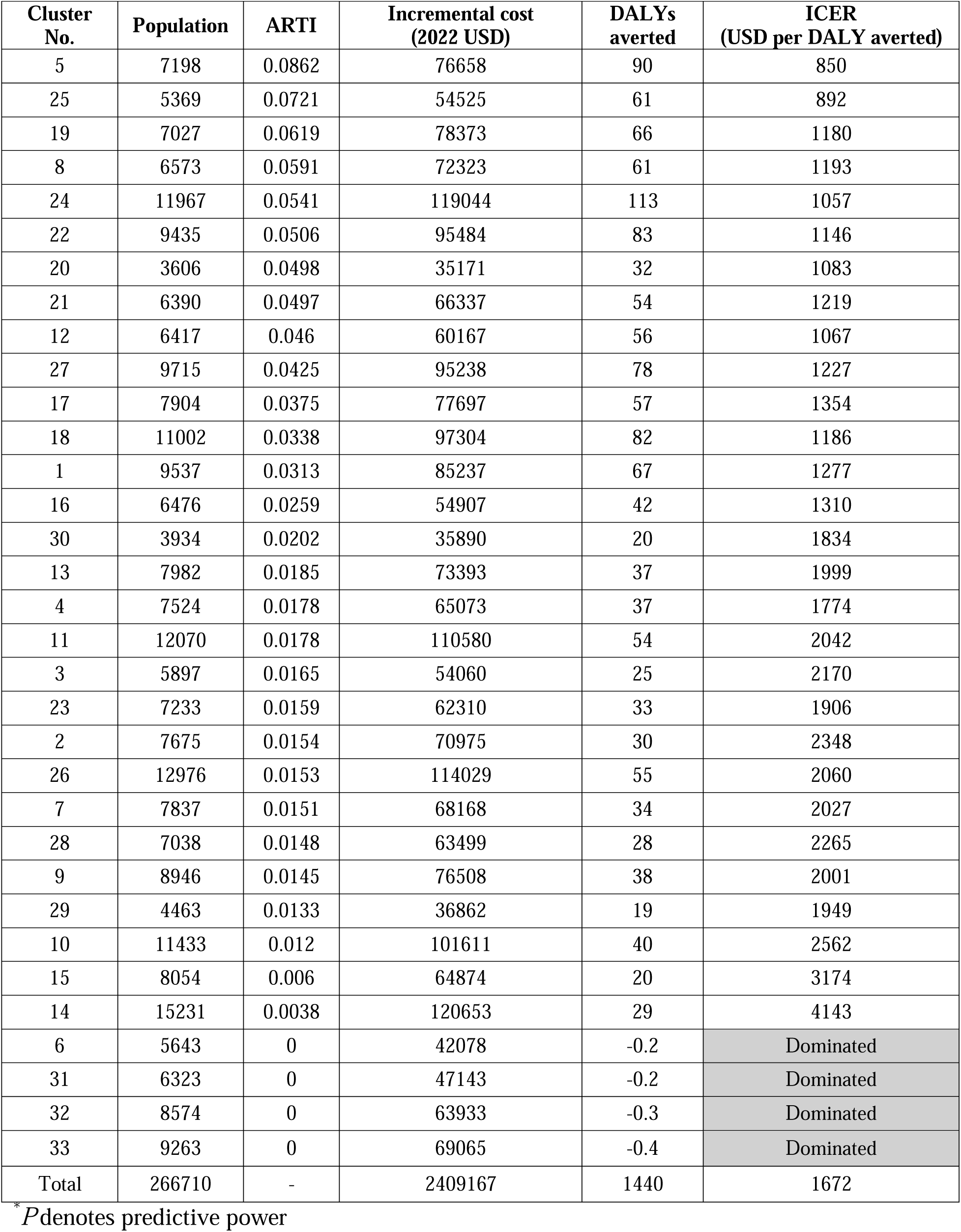
Cost-effectiveness of targeted ACF by cluster (*P*\*=0.6).

| Cluster No. | Population | ARTI | Incremental cost (2022 USD) | DALYs averted | ICER (USD per DALY averted) |
| --- | --- | --- | --- | --- | --- |
| 5 | 7198 | 0.0862 | 76658 | 90 | 850 |
| 25 | 5369 | 0.0721 | 54525 | 61 | 892 |
| 19 | 7027 | 0.0619 | 78373 | 66 | 1180 |
| 8 | 6573 | 0.0591 | 72323 | 61 | 1193 |
| 24 | 11967 | 0.0541 | 119044 | 113 | 1057 |
| 22 | 9435 | 0.0506 | 95484 | 83 | 1146 |
| 20 | 3606 | 0.0498 | 35171 | 32 | 1083 |
| 21 | 6390 | 0.0497 | 66337 | 54 | 1219 |
| 12 | 6417 | 0.046 | 60167 | 56 | 1067 |
| 27 | 9715 | 0.0425 | 95238 | 78 | 1227 |
| 17 | 7904 | 0.0375 | 77697 | 57 | 1354 |
| 18 | 11002 | 0.0338 | 97304 | 82 | 1186 |
| 1 | 9537 | 0.0313 | 85237 | 67 | 1277 |
| 16 | 6476 | 0.0259 | 54907 | 42 | 1310 |
| 30 | 3934 | 0.0202 | 35890 | 20 | 1834 |
| 13 | 7982 | 0.0185 | 73393 | 37 | 1999 |
| 4 | 7524 | 0.0178 | 65073 | 37 | 1774 |
| 11 | 12070 | 0.0178 | 110580 | 54 | 2042 |
| 3 | 5897 | 0.0165 | 54060 | 25 | 2170 |
| 23 | 7233 | 0.0159 | 62310 | 33 | 1906 |
| 2 | 7675 | 0.0154 | 70975 | 30 | 2348 |
| 26 | 12976 | 0.0153 | 114029 | 55 | 2060 |
| 7 | 7837 | 0.0151 | 68168 | 34 | 2027 |
| 28 | 7038 | 0.0148 | 63499 | 28 | 2265 |
| 9 | 8946 | 0.0145 | 76508 | 38 | 2001 |
| 29 | 4463 | 0.0133 | 36862 | 19 | 1949 |
| 10 | 11433 | 0.012 | 101611 | 40 | 2562 |
| 15 | 8054 | 0.006 | 64874 | 20 | 3174 |
| 14 | 15231 | 0.0038 | 120653 | 29 | 4143 |
| 6 | 5643 | 0 | 42078 | -0.2 | Dominated |
| 31 | 6323 | 0 | 47143 | -0.2 | Dominated |
| 32 | 8574 | 0 | 63933 | -0.3 | Dominated |
| 33 | 9263 | 0 | 69065 | -0.4 | Dominated |
| Total | 266710 | - | 2409167 | 1440 | 1672 |

\* $P$ denotes predictive power

**Appendix Table 9:** Cost-effectiveness of targeted ACF by cluster (ARTI *P*\*=0.4).

| Cluster No. | Population | ARTI | Incremental cost (2022 USD) | DALYs averted | ICER (USD per DALY averted) |
| --- | --- | --- | --- | --- | --- |
| 5 | 7198 | 0.0862 | 72291 | 73 | 990 |
| 25 | 5369 | 0.0721 | 52198 | 51 | 1018 |
| 19 | 7027 | 0.0619 | 74879 | 57 | 1303 |
| 8 | 6573 | 0.0591 | 69375 | 53 | 1310 |
| 24 | 11967 | 0.0541 | 115738 | 100 | 1157 |
| 22 | 9435 | 0.0506 | 92998 | 75 | 1239 |
| 20 | 3606 | 0.0498 | 34375 | 29 | 1172 |
| 21 | 6390 | 0.0497 | 64548 | 49 | 1312 |
| 12 | 6417 | 0.046 | 59161 | 52 | 1143 |
| 27 | 9715 | 0.0425 | 93712 | 72 | 1294 |
| 17 | 7904 | 0.0375 | 76885 | 55 | 1401 |
| 18 | 11002 | 0.0338 | 96950 | 80 | 1210 |
| 1 | 9537 | 0.0313 | 85123 | 66 | 1285 |
| 16 | 6476 | 0.0259 | 55107 | 43 | 1276 |
| 30 | 3934 | 0.0202 | 36432 | 21 | 1718 |
| 13 | 7982 | 0.0185 | 74808 | 40 | 1848 |
| 4 | 7524 | 0.0178 | 66065 | 41 | 1621 |
| 11 | 12070 | 0.0178 | 112854 | 60 | 1875 |
| 3 | 5897 | 0.0165 | 55346 | 28 | 1968 |
| 23 | 7233 | 0.0159 | 63448 | 37 | 1707 |
| 2 | 7675 | 0.0154 | 72943 | 35 | 2108 |
| 26 | 12976 | 0.0153 | 116527 | 63 | 1837 |
| 7 | 7837 | 0.0151 | 69605 | 39 | 1801 |
| 28 | 7038 | 0.0148 | 65177 | 32 | 2015 |
| 9 | 8946 | 0.0145 | 78047 | 44 | 1762 |
| 29 | 4463 | 0.0133 | 37497 | 22 | 1682 |
| 10 | 11433 | 0.012 | 104910 | 48 | 2197 |
| 15 | 8054 | 0.006 | 66708 | 28 | 2355 |
| 14 | 15231 | 0.0038 | 124285 | 44 | 2803 |
| 6 | 5643 | 0 | 42078 | -0.2 | Dominated |
| 31 | 6323 | 0 | 47143 | -0.2 | Dominated |
| 32 | 8574 | 0 | 63933 | -0.3 | Dominated |
| 33 | 9263 | 0 | 69065 | -0.4 | Dominated |
| Total | 266710 | - | 2410212 | 1440 | 1674 |

\* $P$ denotes predictive power

**Appendix Table 10:** Cost-effectiveness of targeted ACF by cluster (ARTI *P*\*=0.2).

| Cluster No. | Population | ARTI | Incremental cost (2022 USD) | DALYs averted | ICER (USD per DALY averted) |
| --- | --- | --- | --- | --- | --- |
| 5 | 7198 | 0.0862 | 68442 | 58 | 1183 |
| 25 | 5369 | 0.0721 | 50034 | 42 | 1188 |
| 19 | 7027 | 0.0619 | 71452 | 49 | 1469 |
| 8 | 6573 | 0.0591 | 66431 | 45 | 1468 |
| 24 | 11967 | 0.0541 | 112311 | 87 | 1290 |
| 22 | 9435 | 0.0506 | 90336 | 66 | 1365 |
| 20 | 3606 | 0.0498 | 33516 | 26 | 1291 |
| 21 | 6390 | 0.0497 | 62615 | 44 | 1438 |
| 12 | 6417 | 0.046 | 58023 | 47 | 1247 |
| 27 | 9715 | 0.0425 | 91878 | 66 | 1390 |
| 17 | 7904 | 0.0375 | 75754 | 51 | 1473 |
| 18 | 11002 | 0.0338 | 96303 | 77 | 1257 |
| 1 | 9537 | 0.0313 | 84723 | 64 | 1317 |
| 16 | 6476 | 0.0259 | 55168 | 44 | 1266 |
| 30 | 3934 | 0.0202 | 36861 | 22 | 1639 |
| 13 | 7982 | 0.0185 | 76018 | 44 | 1739 |
| 4 | 7524 | 0.0178 | 66937 | 44 | 1509 |
| 11 | 12070 | 0.0178 | 114855 | 66 | 1753 |
| 3 | 5897 | 0.0165 | 56531 | 31 | 1819 |
| 23 | 7233 | 0.0159 | 64517 | 41 | 1559 |
| 2 | 7675 | 0.0154 | 74820 | 39 | 1930 |
| 26 | 12976 | 0.0153 | 118919 | 71 | 1671 |
| 7 | 7837 | 0.0151 | 70989 | 43 | 1633 |
| 28 | 7038 | 0.0148 | 66808 | 37 | 1829 |
| 9 | 8946 | 0.0145 | 79556 | 50 | 1584 |
| 29 | 4463 | 0.0133 | 38141 | 26 | 1483 |
| 10 | 11433 | 0.012 | 108386 | 56 | 1926 |
| 15 | 8054 | 0.006 | 69048 | 38 | 1798 |
| 14 | 15231 | 0.0038 | 129442 | 66 | 1963 |
| 6 | 5643 | 0 | 42078 | -0.2 | Dominated |
| 31 | 6323 | 0 | 47143 | -0.2 | Dominated |
| 32 | 8574 | 0 | 63933 | -0.3 | Dominated |
| 33 | 9263 | 0 | 69065 | -0.4 | Dominated |
| Total | 266710 | - | 2411033 | 1439 | 1675 |

\* $P$ denotes predictive power

**Appendix Table 11:** Cost-effectiveness of untargeted ACF by cluster (ARTI *P*\*=0).

| Cluster No. | Population | ARTI | Incremental cost (2022 USD) | DALYs averted | ICER (USD per DALY averted) |
| --- | --- | --- | --- | --- | --- |
| 5 | 7198 | 0.0862 | 63847 | 40 | 1606 |
| 25 | 5369 | 0.0721 | 47169 | 30 | 1573 |
| 19 | 7027 | 0.0619 | 66422 | 36 | 1859 |
| 8 | 6573 | 0.0591 | 61948 | 34 | 1846 |
| 24 | 11967 | 0.0541 | 106680 | 66 | 1624 |
| 22 | 9435 | 0.0506 | 85667 | 51 | 1692 |
| 20 | 3606 | 0.0498 | 31980 | 20 | 1606 |
| 21 | 6390 | 0.0497 | 59158 | 33 | 1770 |
| 12 | 6417 | 0.046 | 55796 | 36 | 1537 |
| 27 | 9715 | 0.0425 | 87863 | 52 | 1677 |
| 17 | 7904 | 0.0375 | 72614 | 42 | 1738 |
| 18 | 11002 | 0.0338 | 93883 | 64 | 1477 |
| 1 | 9537 | 0.0313 | 82589 | 54 | 1524 |
| 16 | 6476 | 0.0259 | 54313 | 38 | 1424 |
| 30 | 3934 | 0.0202 | 36266 | 21 | 1753 |
| 13 | 7982 | 0.0185 | 74977 | 41 | 1832 |
| 4 | 7524 | 0.0178 | 66335 | 42 | 1585 |
| 11 | 12070 | 0.0178 | 113469 | 62 | 1835 |
| 3 | 5897 | 0.0165 | 56013 | 30 | 1881 |
| 23 | 7233 | 0.0159 | 64154 | 40 | 1606 |
| 2 | 7675 | 0.0154 | 74327 | 38 | 1973 |
| 26 | 12976 | 0.0153 | 118328 | 69 | 1709 |
| 7 | 7837 | 0.0151 | 70686 | 42 | 1667 |
| 28 | 7038 | 0.0148 | 66517 | 36 | 1859 |
| 9 | 8946 | 0.0145 | 79345 | 49 | 1606 |
| 29 | 4463 | 0.0133 | 38147 | 26 | 1482 |
| 10 | 11433 | 0.012 | 108889 | 58 | 1894 |
| 15 | 8054 | 0.006 | 70612 | 45 | 1565 |
| 14 | 15231 | 0.0038 | 133985 | 85 | 1577 |
| 6 | 5643 | 0 | 50600 | 31 | 1645 |
| 31 | 6323 | 0 | 61095 | 31 | 1962 |
| 32 | 8574 | 0 | 79502 | 45 | 1776 |
| 33 | 9263 | 0 | 88373 | 46 | 1902 |
| Total | 266710 | - | 2421550 | 1431 | 1692 |

\* $P$ denotes predictive power

